# Preference for Blood-Based Colorectal Cancer Screening in the Black Community

**DOI:** 10.64898/2026.07.22.26358691

**Authors:** R. Li, T.M. Jackson, J. Onyekaba, Y. He, K. Njoku, C. Amadi, K. Chandora, D. Ortega, H. Gundroo, T. Tobun, C. Smith, D. Hommes, J.M. Luévano, J.J. Liu

## Abstract

**Background:** Blood-based colorectal cancer (CRC) screening is a novel, less invasive screening that has the potential to improve CRC screening adherence, particularly among Black American populations, which has historically lower screening rates. We evaluated factors associated with preference for blood-based CRC screening among Black adults in church-based community settings.

**Methods:** From October 2023 to January 2024, a cross-sectional survey was conducted over 101 adults aged 45 to 75 years at three Black churches in metropolitan Atlanta, Georgia. Demographics, CRC screening history, healthcare access, and attitudes toward blood-based CRC screening were assessed.

**Results:** Overall, 72 participants (71.3%) preferred blood-based CRC screening. Preference was not associated with demographic or socioeconomic characteristics, screening history, or healthcare access. The most commonly reported reasons were ease of testing (83.3%), avoidance of stool collection (38.9%), avoidance of bowel preparation (36.1%), perceived fewer side effects (34.7%), and perceived lower risk (33.3%). Needle concern was the only factor significantly associated with non-preference (p=0.034).

**Conclusion:** Blood-based colorectal cancer screening was preferred by most Black adults and may improve CRC screening participation through greater convenience.

## Introduction

Colorectal cancer (CRC) is largely preventable with routine screening but remains the second leading cause of cancer-related mortality in the United States.^1^ Black Americans, in particular, experience disproportionally higher incidence and mortality rates than other racial and ethnic groups.^2^ Socioeconomic factors, access to preventive care, and lifestyle habits may all contribute to disparities in screening participation.^2,3^ To identify barriers to CRC screening and design effective interventions, it is important to understand Black patients’ knowledge, attitudes, and preferences regarding CRC screening modalities.^4^ Understanding patient preferences has become increasingly important with emergence of blood-based tests, which can be readily integrated into routine clinical and may improve CRC screening rates.^5,6^

Regular screening has led to substantial declines in CRC incidence and mortality, yet around a third of eligible Americans remain unscreened, and estimated 46–63% of CRC deaths are attributed to missed screening opportunities.^1,3,5,7^ Although stool-based testing and colonoscopy are effective screening methods^3,5,7,9^, their real-world benefits are limited by poor adherence and acceptability.^3,8^ Colonoscopy remains the gold standard for CRC screening but requires bowel preparation and an invasive procedure associated with discomfort and complication risks.^3,9^ Stool-based tests are noninvasive and convenient but require stool-handling and repeated testing, which many patients find undesirable.^10,11,14^

The recently FDA-approved blood-based CRC screening test, Shield (Guardant, Palo Alto, CA) offers a new screening option that can be readily integrated into routine clinical care and may help address several barriers associated with conventional CRC screening.^5,12,13^ Its main limitations include lower sensitivity for advanced adenomas and the need for follow-up diagnostic colonoscopy after a positive result.^12,13^ Furthermore, evidence regarding its long-term effectiveness in reducing CRC incidence and mortality is still evolving.^3,13^ Given these tradeoffs, understanding Black patients’ preferences for blood-based CRC screening is essential to guide implementation of this emerging screening modality in a population disproportionately affected by CRC.^2,4^ We conducted this study to identify factors associated with preference for blood-based CRC screening among Black Americans in a community setting.

## Methods

### Survey

This descriptive, cross-sectional study was conducted via a self-administered survey from October 2023 to January 2024 in collaboration with three Black churches in metropolitan areas in Atlanta, Georgia. The survey collected information on demographics, healthcare access, CRC screening history, screening preferences, and attitudes toward blood-based CRC screening.

### Participants

Eligible participants were adults aged 45–75 years who attended church services and met the recommended age for average-risk CRC screening. Individuals at high risk for CRC, including those with inflammatory bowel disease, hereditary colorectal cancer syndromes, or cystic fibrosis, were excluded.

### Statistical Analysis

Descriptive statistics summarized participant characteristics. Categorical variables were reported as frequencies and percentages. Fisher’s exact test was used for group comparisons, with p<0.05 considered statistically significant. Analyses were performed using STATA version 18.5 (StataCorp, College Station, TX). This study was approved by the Institutional Review Board at Morehouse School of Medicine.

## Results

A total of 101 participants completed the survey, 72 (71.3%) participants responded “Yes” to blood-based screening preferences, while 29 (28.7%) answered “No.” The mean age was 64.3 +/-9.6 years among participants who preferred blood-based screening and 65.8 +/-8.3 years among those who did not. Most participants were Black (N=97, 96%) who had primary care provider (N=95, 94%) and trusted their primary care provider (N=92, 91%).

Demographic and socioeconomic characteristics, including age, sex, race, ethnicity, marital status, education, employment status, insurance status, and income, were not associated with preference for blood-based screening (Table 1). Similarly, CRC screening history, intention to undergo screening in the next year, and reported barriers to conventional CRC screening did not differ by screening preference. A personal history of colorectal polyps was the only clinical characteristic significantly associated with preference for blood-based screening (P = 0.0152).

**Table 1:**
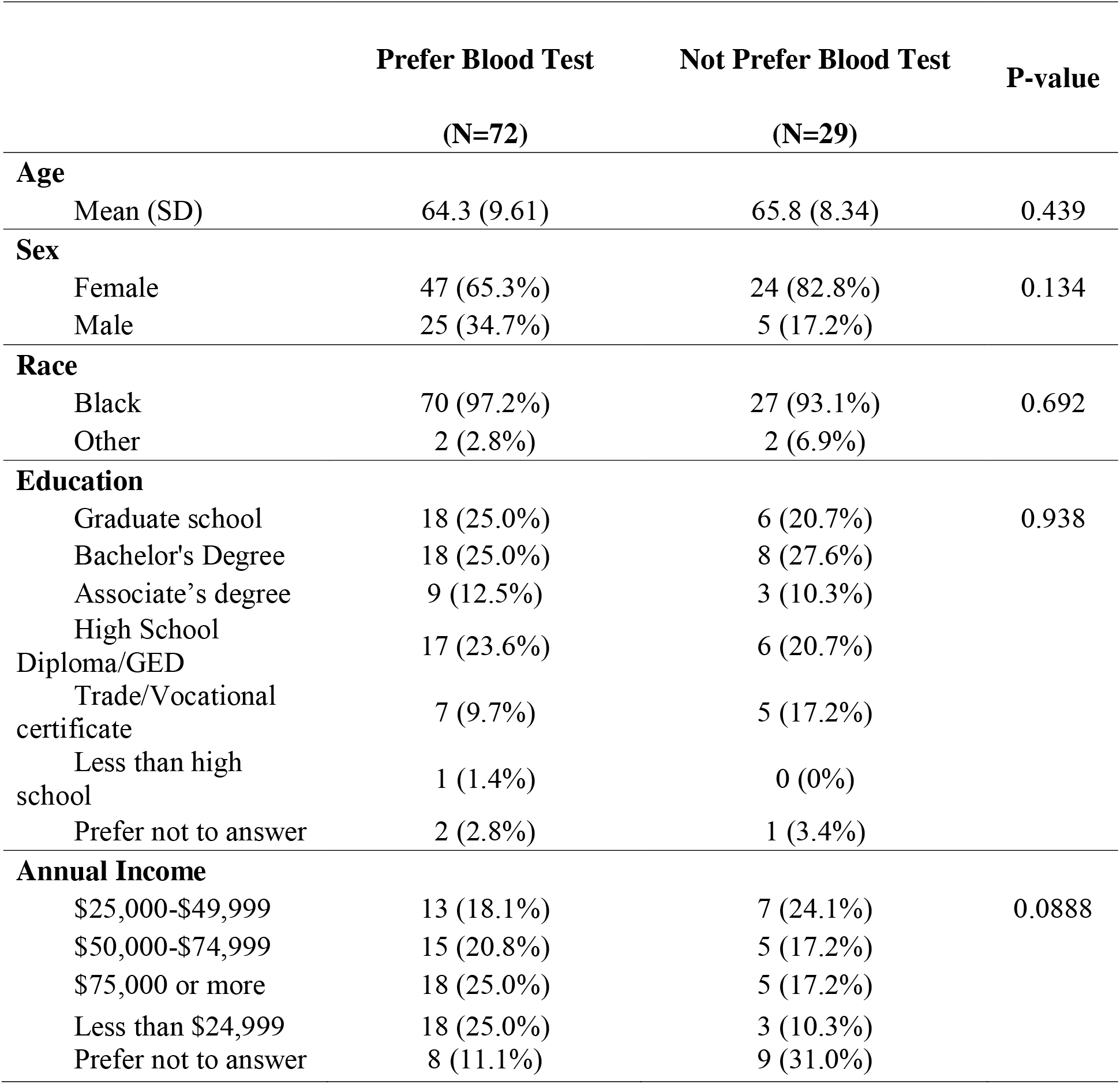
Demographic and Social Economic Characteristics.

Among participants who preferred blood-based screening, the most frequently cited reason was ease of testing (83.3%). Other reasons included avoiding stool collection (38.9%), avoiding fasting or bowel preparation (36.1%), perceived fewer side effects (34.7%), perceived lower risk (33.3%), and avoiding time away from work (22.2%), shown in Table 2.

**Table 2.** Factors Influencing Blood-Based Colorectal Cancer Screening.

|  | Prefer Blood<br>Screening:<br>(N=72) | Not Prefer Blood<br>Screening:<br>(N=29) | P-value |
| --- | --- | --- | --- |
| Private Insurance | 27 (37.5%) | 7 (24.1%) | 0.292 |
| Family Polyps History | 12 (16.7%) | 2 (6.9%) | 0.139 |
| Family CRC History | 14 (19.4%) | 5 (17.2%) | 0.762 |
| Personal Polyp History | 20 (27.8%) | 5 (17.2%) | 0.0152* |
| Personal CRC History | 3 (4.2%) | 1 (3.4%) | 0.283 |
| Have PCP | 67 (93.1%) | 28 (96.6%) | 0.836 |
| Trust PCP | 66 (91.7%) | 26 (89.7%) | 0.841 |
| PCP Discussing Screening Options | 48 (66.7%) | 17 (58.6%) | 0.692 |
| Reasons for preferring blood-based screening |  |  |  |
| Ease of testing | 60 (83.3%) | 0 (0%) | <0.001* |
| Perceived Lower Risk | 24 (33.3%) | 0 (0%) | <0.001* |
| Fewer Side effects | 25 (34.7%) | 0 (0%) | <0.001* |
| Avoid Bowel Preparation | 26 (36.1%) | 0 (0%) | <0.001* |
| Avoid Stool Collection | 28 (38.9%) | 0 (0%) | <0.001* |
| No Time Off Work | 16 (22.2%) | 0 (0%) | 0.0137* |
| Reasons for not preferring blood-based screening |  |  |  |
| Needle Concerns | 0 (0%) | 3 (10.3%) | 0.0338* |
| Uncomfortable Sharing Blood Samples | 0 (0%) | 2 (6.9%) | 0.144 |
| Need for a Clinic Visit | 0 (0%) | 1 (3.4%) | 0.636 |
| Medical Mistrust | 0 (0%) | 1 (3.4%) | 0.636 |

Among participants who did not prefer blood-based screening, concerns about needles 10.3% was the only factor associated with non-preference (P = 0.034). Other concerns such as discomfort with sharing blood, the need for a clinic visit, and mistrust were not significantly associated with blood-based screening preference.

## Discussion

In our study, we found that blood-based screening test was preferred by majority (71.3%) of Black patients in this church community-based cohort. Preference for blood-based screening was not significantly associated with socioeconomic, educational, or insurance-related factors, suggesting that its appeal may extend across demographic differences.

The high rate of preference for blood-based CRC screening observed in our study is consistent with previous reports demonstrating strong patient interest in blood-based screening, particularly among individuals who had previously declined screening colonoscopy.^14^ Similar findings have also been reported in national surveys evaluating patient preferences for CRC screening modalities.^4^ Although these studies were conducted in different populations and healthcare settings, they consistently identified convenience-related factors as important determinants of screening preference. Likewise, participants in our study most often cited ease of testing, avoidance of stool collection, avoidance of bowel preparation, and perceived lower risk and fewer side effects as reasons for preferring blood-based screening, consistent with previous reports of patient preference for less invasive screening options.^6,11^ The strong preference for blood-based screening observed in our study suggests that convenience-related factors may play an important role in CRC screening decisions among Black adults.

Although participants who preferred blood-based testing readily cited specific reasons for their choice, few participants in the non-preference group (N = 29) identified reasons against it. This asymmetry suggests that the factors driving non-preference may be more complex or less immediately accessible than those motivating preference and may not be fully captured by the closed-ended survey items used in this study. Future studies incorporating qualitative interviews or open-ended survey questions may provide a more comprehensive understanding of the factors influencing non-preference for blood-based CRC screening.

Our study has several limitations. First, the relatively small sample size limited statistical power, particularly for identifying factors associated with non-preference for blood-based screening. Second, participants were recruited from only three Black metropolitan churches in one city in the South (Atlanta, Georgia), which may limit the generalizability of our findings to Black Americans in other geographic areas or those not engaged in faith-based communities. Third, the cross-sectional design precludes causal inference and cannot determine whether stated preferences translate into actual screening completion and adherence. Finally, our survey only included limited range of potential reasons for screening preference and non-preference, which may have provided an incomplete picture of the factors influencing participants’ decisions.

In conclusion, we found a high rate of preference for blood-based CRC screening among Black adults in community-based settings, with 71.3% of participants preferring this screening modality. Convenience of blood collection, avoidance of stool handling and bowel preparation, and perceived lower procedural risk were the primary factors influencing this preference. These findings may help lead patient-centered strategies to improve CRC screening rates among Black communities.^2^

## Data Availability

All data produced in the present study are available upon reasonable request to the authors.

